# Beyond one-off training: Continuing support, system stability and worker-borne costs shape electronic community health reporting in Kisii County, Kenya

**DOI:** 10.64898/2026.11.10.26367703

**Authors:** Maureen Atieno Adoyo, Eunice Bitutu Koringo

**Author notes:** Corresponding author: Maureen Adoyo, (MA). MAA & EBK contributed equally to this work.

## Abstract

Kenya’s electronic community health information system (eCHIS) moves household and service data from community health promoters’ phones into the national health information system. When reporting quality falls short, the usual response is more training and competency frameworks. We tested whether user capability and one-off training explain how well the system performs for its users, or whether continuing support and system stability matter more. We also examined where quality was lost along the reporting chain. In this convergent mixed-methods cross-sectional study in Kisii County, we surveyed 343 eCHIS users in 22 facility catchments, observed five sites against a structured checklist, and interviewed 13 key informants. Hierarchical ordinal and logistic regression with facility-clustered standard errors modelled user satisfaction and perceived system consistency. Formal eCHIS training was near universal (94.5%) and was not associated with either outcome. Adding continuing support (refresher training, mentorship and technical support response time) significantly improved both models. McFadden’s pseudo-R² for consistency rose from 0.06 to 0.16. Each faster level of technical support response almost doubled the odds of reporting a consistent system (adjusted odds ratio 1.87, 95% CI 1.35-2.58). Each step up in crash frequency reduced the odds of higher satisfaction (0.60, 0.43-0.84). Digital comfort remained independently associated with both outcomes. At the observation sites, form logic secured completeness (all required fields were filled at 5 of 5 sites), but timeliness (2 of 5), consistency over time (1 of 5) and use of reports for decisions (0 of 4) failed, mostly because of synchronisation. In open-ended responses, 43.4% of respondents raised the cost of data bundles. Informants described promoters paying for connectivity and reinstalling the failing application themselves. Reporting quality depended less on what users knew than on the support around them. Competency frameworks will need matching standards for connectivity financing, support response and synchronisation monitoring.

**Author Summary:** Community health promoters in Kenya now record household visits on smartphones instead of paper. The aim is faster, more accurate data for planning. When the data disappoint, the usual answer is to train the promoters again. We wanted to know whether that answer fits the problem. We surveyed 343 system users in Kisii County, visited five health sites to watch the system in use, and interviewed 13 supervisors and managers. Nearly everyone had been trained, and training made no difference to how well the system worked for them. What did make a difference was how quickly technical help arrived, how often the application crashed, and whether users had ongoing refresher training and mentoring. At the sites we visited, the application made sure every form was complete. The problems came later, when records had to be uploaded over weak networks using data bundles that promoters often paid for themselves. Reports were late, sometimes duplicated, and not used for decisions. We conclude that improving community health data in Kenya means paying for connectivity, responding quickly to system failures and watching whether data actually arrive, not only training workers again.

## 1. Introduction

Community health workers are often the only contact that rural households have with the formal health system, and the records they keep are the main source of information on what happens at that level. Electronic community health information systems were introduced to replace paper registers with mobile applications. These applications capture household and service data at the point of care and pass them to national reporting platforms. Reviews of community-based health information systems in low- and middle-income countries show that digitisation can improve completeness and timeliness through mandatory fields, automated validation and real-time dashboards. The same reviews show that these gains are fragile, uneven, and dependent on the conditions in which the systems are run.^1–4^

Kenya has moved quickly in this direction. A national electronic community health information system (eCHIS) was co-created with government and partners and rolled out to community health promoters (CHPs) county by county ^5^. Early Kenyan evaluations report better timeliness and accessibility of community data. They also report barriers that recur across settings: unreliable connectivity, system downtime, thin technical support and training that is short and seldom repeated ^6^. In Homa Bay County, stakeholders described network instability as the main obstacle to using digital health data for care and decisions ^7^. National-level users of the District Health Information Software 2 (DHIS2) question the accuracy of routine data ^8^. Similar patterns appear in Ethiopia, where eCHIS was adopted earlier and has been studied more closely^9–12^

The dominant policy response to weak data quality has been to treat it as a problem of the workforce. Training curricula, digital literacy programmes and, increasingly, national competency frameworks are proposed to close the gap. The evidence for this response is narrower than its popularity suggests. Training reliably improves knowledge, confidence and digital skills among community health workers ^13,14^ However, the interventions that have improved routine data quality in controlled or implementation studies bundled training with supportive supervision, mentorship, feedback and recognition, and none isolated training as the active ingredient ^15,16^. Ethiopian eCHIS users described their basic training as too short and too theoretical, with little follow-up ^9^. Acceptance of the system was close to universal, but actual use was about half that level ^17^. Studies that measured routine data quality directly tend to link it to refresher training, supervision and user-friendly tools rather than to initial training alone ^18,19^

A second body of evidence locates the problem in infrastructure. In sub-Saharan Africa, unstable connectivity, power interruptions and failed synchronisation delay the movement of records from the point of capture to central systems. They also create duplicate, missing and conflicting records.^20–24^ Offline-first design protects data capture during outages but does not restore real-time availability, which still depends on later successful synchronization. ^25,26^ Systems that performed well under difficult conditions did so because they combined offline capability with validation, device management and intensive technical support ^26,27^. A third strand shows who pays to keep these systems running. Community health workers frequently absorb the recurrent costs of digital work, including airtime, mobile data, charging and device repair ^28–31^. When these costs are not covered, motivation and performance suffer, and retention also declines ^32–34^

Sociotechnical frameworks bring these strands together. The Performance of Routine Information System Management (PRISM) framework separates technical, organisational and behavioural determinants of information system performance ^35,36^. The design-reality gap perspective explains failure as a mismatch between the conditions a system assumes and those it meets ^37^ More recent infrastructure-aware approaches treat power and connectivity as determinants that shape the whole system, not as background nuisances ^25,38,39^. Taken together, these perspectives imply that the quality of a community data stream depends on where along the reporting chain it is examined. At the point of capture, the application’s own form logic can enforce completeness regardless of the setting. Synchronisation, timely arrival, concordance with source records and use for decisions are different. They depend on a support ecology of connectivity, power, recurrent financing, technical response and continuing learning that the application does not control (Fig 1).

**Fig 1.**
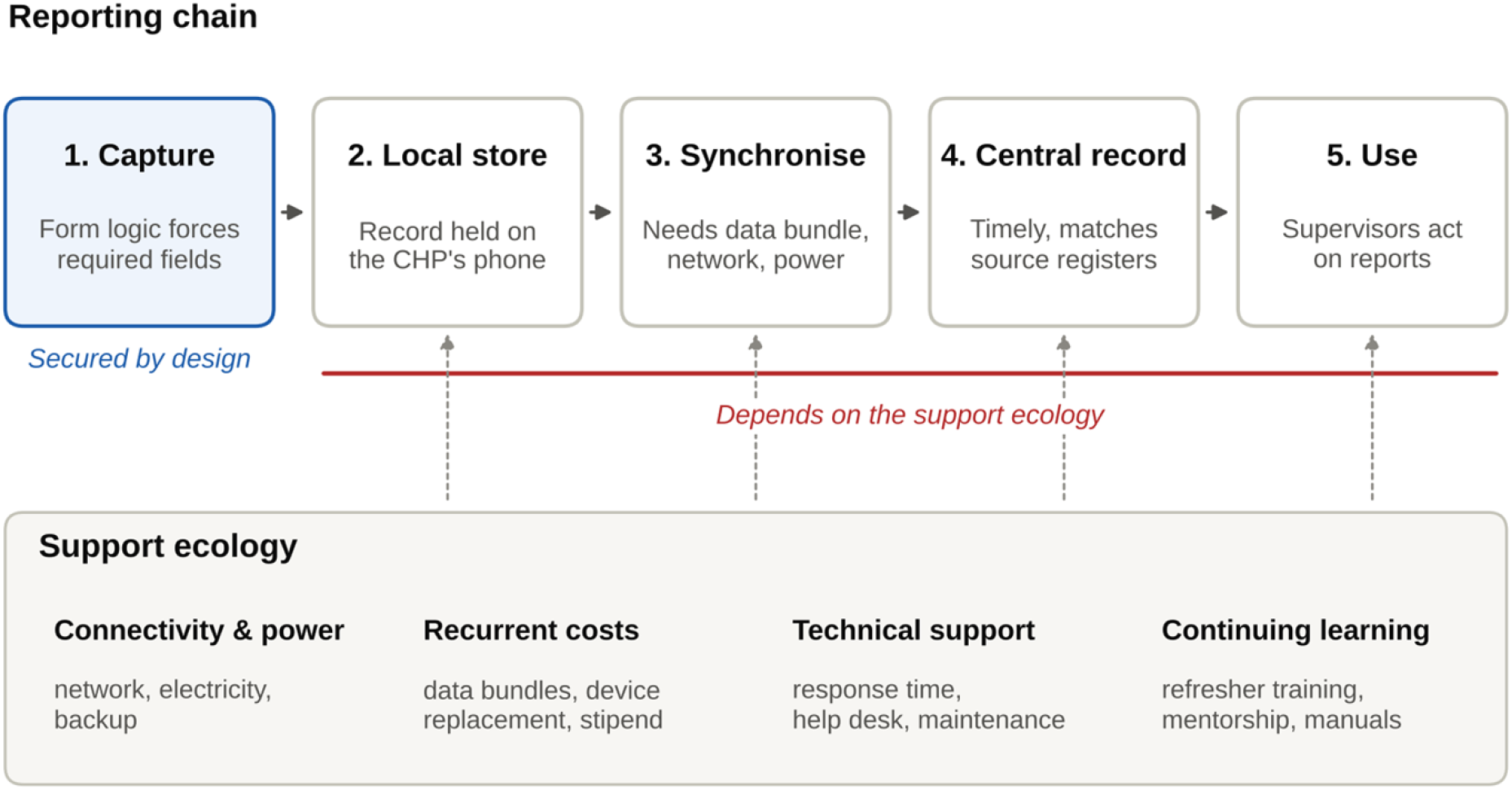
The reporting chain and its support ecology. Conceptual model used in this study. The application’s form logic secures completeness at the point of capture. Later stages depend on connectivity, power, recurrent financing, technical support and continuing learning, which the application does not control.

Few studies have tested these competing explanations in the same population, and none, to our knowledge, in a Kenyan county using the national eCHIS. In this study we used survey, observation and interview data from Kisii County to answer four questions. First, is one-off formal training associated with how well the system performs for its users? Second, do continuing support and system performance add explanatory value beyond users’ own digital capability? Third, at which stages of the reporting chain is data quality secured or lost? Fourth, who carries the costs of keeping the system running? We argue that, in this setting, reporting quality depended less on what users knew than on the support around them, and that competency-centred reforms will not close the gap on their own.

## 2. Materials and methods

### 2.1 Study design and setting

We conducted a cross-sectional study using a convergent parallel mixed-methods design. We administered a structured survey, a site observation checklist, and key informant interviews during the same period and integrated them at the interpretation stage ^40^. Reporting of the quantitative component follows the STROBE guidelines for cross-sectional studies ^40^.

Kisii County lies in western Kenya and comprises nine sub-counties. CHPs deliver community health services from community units, each linked to a public health facility. Community health assistants (CHAs) supervise CHPs, and sub-county and county community health focal persons coordinate the programme. CHPs use the eCHIS application on government-issued smartphones to register households, record services and referrals, and submit routine reports. These data are synchronised with the national system. According to county programme records, the eligible workforce totalled 3,047: 2,940 CHPs, 97 CHAs, nine sub-county focal persons and one county focal person.

### 2.2 Participants and sampling

The sample size was calculated with Cochran’s formula for a proportion (95% confidence, 5% precision, p = 0.5)^41^. With a finite population correction for 3,047 and a 5% allowance for non-response, the target was 359. The sample was stratified by sub-county and cadre, and respondents were selected by simple random sampling within strata. Eligible respondents had used eCHIS in their routine work, were available during data collection and gave written informed consent. Key informants were selected purposively for their supervisory or coordinating roles. Observation sites were selected purposively to cover five sub-counties and three levels of facility.

### 2.3 Instruments and data collection

The questionnaire contained 27 items. They covered respondent characteristics; eCHIS training and its frequency; mentorship; digital comfort and prior digital experience; technical support availability and response time; frequency of system crashes; ease of access; device used; perceived system consistency; satisfaction; staff attrition and retention; and data privacy. Most items had ordered response categories. Three items allowed multiple or free-text responses: usage challenges, areas needing improvement, and recommendations. The questionnaire was pretested with 18 CHPs and its wording simplified where items proved hard to understand.

The observation checklist contained 40 yes/no indicators with space for comments, grouped into five domains: support infrastructure, user conditions, competence, system maintenance and quality of reporting. Three trained observers completed it at five sites (three hospitals, one health centre and one dispensary) between 22 September and 27 October 2025. The key informant guide covered the same domains. We administered it to 13 informants: 11 CHAs, one sub-county focal person, and one county focal person. We recorded responses as structured summaries against each question in a response template.

### 2.4 Variables

The survey did not directly measure data quality. We therefore analysed two outcomes that capture how well the system performed for its users, and we used the observation and interview data to assess data quality along the reporting chain. The first outcome was satisfaction with eCHIS on a five-point ordinal scale from very dissatisfied to very satisfied. The second was perceived system consistency, a binary response to whether the system worked consistently. Respondents who answered “not sure” (n = 30) were excluded from the main analysis of this outcome.

We grouped explanatory variables into four conceptual blocks. User capability comprised digital comfort (three levels), prior digital experience (yes/no) and more than six years in community health work (yes/no). One-off training was a single indicator: whether the respondent had ever received formal eCHIS training. Continuing support comprised refresher training frequency (never, annually, quarterly or monthly), availability of a mentorship programme (yes/no) and technical support response time (no support received, more than two days, within a day or immediately). System performance comprised crash frequency (never, once, two to three times or more than three times) and ease of access (four levels). Ordered predictors were entered as linear scores, so each odds ratio refers to a one-level change.

### 2.5 Statistical analysis

Descriptive statistics are reported as counts and percentages of all 343 respondents, with Wilson 95% confidence intervals. We examined bivariate associations using Spearman’s rank correlation. Satisfaction was modelled with proportional-odds ordinal logistic regression^42^, consistent with binary logistic regression. We entered the four blocks in sequence (capability, then one-off training, then continuing support, then system performance). We assessed each block’s contribution using likelihood-ratio tests, McFadden’s pseudo-R², and the Akaike information criterion. Standard errors were clustered on the 22 health facilities. Models used complete cases (n = 307 for satisfaction and n = 281 for consistency). The largest variance inflation factor among predictors was 1.40.

The proportional-odds assumption was examined by fitting separate binary logits at each cut-point. We ran four sensitivity analyses: restricting the sample to CHPs, recoding “not sure” responses as “no”, replacing technical support response time with technical support availability, and dichotomising satisfaction (satisfied or very satisfied versus the rest). For the consistency model, we report average predicted probabilities across levels of support response time, with all other covariates held at their observed values. Analyses were run in Python 3.13 with statsmodels 0.15 and SciPy 1.18. The analysis code is available with the dataset.

### 2.6 Qualitative and observational analysis

Responses to the three multiple-response survey items were coded with a keyword dictionary into ten categories. Each category’s frequency is reported as a share of respondents who answered the item. Observation indicators were tallied as the number of sites meeting each criterion, and observer comments were retained verbatim. Key informant summaries were coded deductively against the PRISM technical, organisational and behavioural domains, with codes added inductively where responses did not fit ^35,43^. We report how many informants expressed each code. The three strands were integrated in a joint display that sets out convergence and divergence for each theme ^44 44^.

### 2.7 Ethics statement

The Institutional Scientific and Ethics Review Committee approved the study (reference number ISERC/KTRH/080/25). Kenya’s National Commission for Science, Technology and Innovation issued a research licence (reference number 876354), and the County Health Research Committee authorised the study. All participants provided written informed consent. No names or other personal identifiers were collected, and facility managers consented to the observations.

## 3. Results

### 3.1 Participants

The dataset contained 343 completed questionnaires, a response rate of 95.5% against the target of 359. Respondents came from 22 facility catchments in all nine sub-counties (Table 1). Nearly all respondents were CHPs (95.9%), and most had worked in community health for more than six years (70.3%). Almost all used a mobile phone to access eCHIS (97.4%).

**Table 1.** Characteristics of survey respondents (n = 343).

| **Characteristic** | **n** | **%** |
|---|---|---|
| **Role** |  |  |
| Community health promoter | 329 | 95.9 |
| Nurse | 4 | 1.2 |
| Data officer | 1 | 0.3 |
| Other | 9 | 2.6 |
| **Years in community health work** |  |  |
| <1 year | 5 | 1.5 |
| 1-3 years | 34 | 9.9 |
| 4-6 years | 63 | 18.4 |
| >6 years | 241 | 70.3 |
| **Device used to access eCHIS** |  |  |
| Mobile phone | 334 | 97.4 |
| Tablet | 7 | 2.0 |
| Laptop | 2 | 0.6 |
| **Sub-county** |  |  |
| Bobasi | 38 | 11.1 |
| Bomachoge Borabu | 38 | 11.1 |
| Bomachoge Chache | 38 | 11.1 |
| Bonchari | 38 | 11.1 |
| Kitutu Chache North | 38 | 11.1 |
| Kitutu Chache South | 39 | 11.4 |
| Nyaribari Chache | 38 | 11.1 |
| Nyaribari Masaba | 38 | 11.1 |
| South Mugirango | 38 | 11.1 |

Respondents were drawn from 22 health facility catchments.

### 3.2 Training, support and system performance

Formal eCHIS training was close to universal (94.5%). Continuing training was not. Only 42.0% received refresher training monthly, 24.8% annually, and 19.0% never. Among the 324 respondents who had been formally trained, 138 (42.6%) had received refresher training no more than once a year, and 58 (17.9%) never had any (Table 2). Digital comfort was high (58.0% very comfortable, 37.3% somewhat comfortable). Technical support was the weakest part of the system. A quarter of respondents (25.1%) had never received support when they needed it, and a further 32.1% had waited more than two days. Together, 57.1% had experienced delayed or absent support. Crashes were common: 70.0% reported at least two and 45.8% more than three. Only 28.3% described the system as consistent, while 63.0% said it was not (Fig 2).

**Fig 2.**
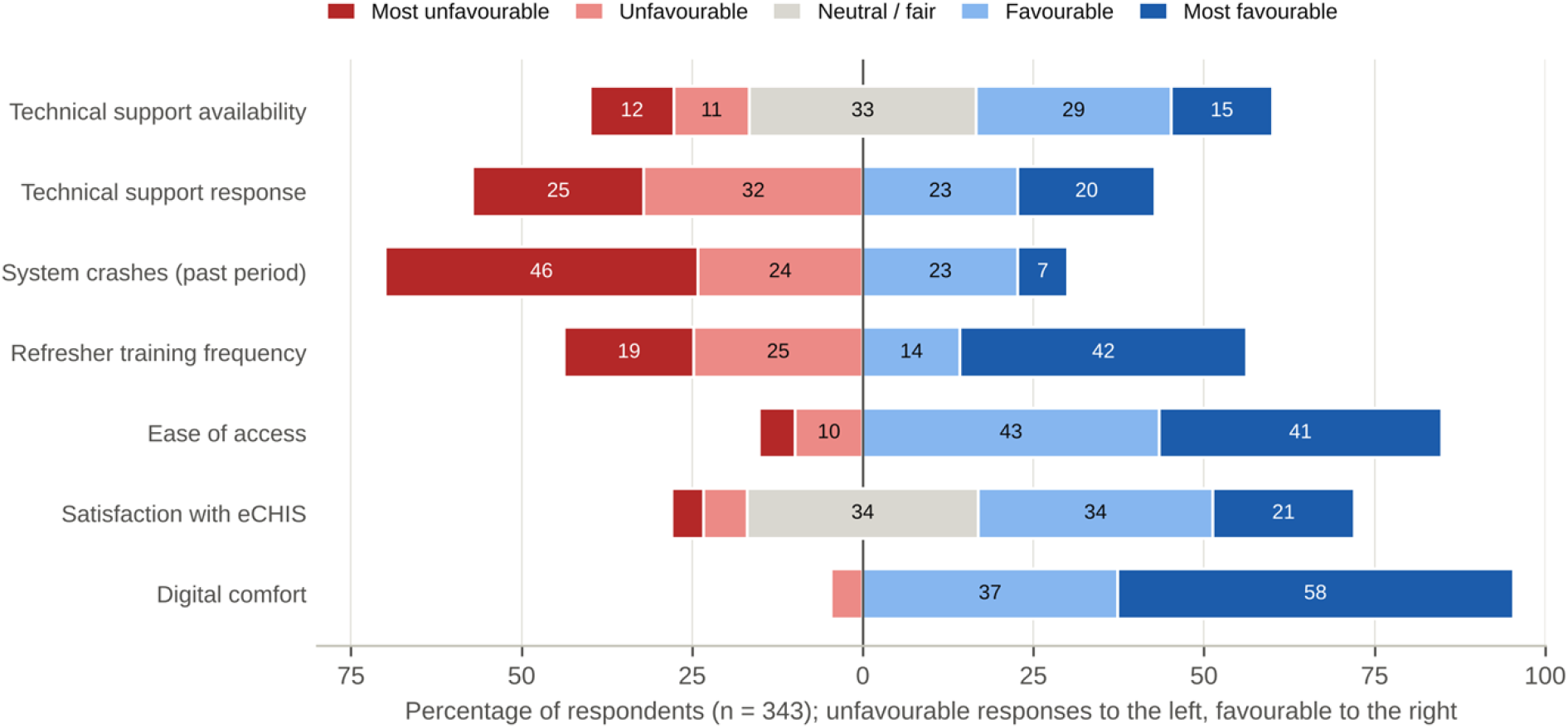
Support, system performance and capability indicators among survey respondents (n = 343). Bars are centred on the neutral category where one exists. Unfavourable responses extend to the left and favourable responses to the right. Values are percentages, and segments below 7% are unlabelled.

**Table 2.** Training, support, system performance and outcome indicators (n = 343).

| Indicator | Response | n | % (95% CI) |
|---|---|---|---|
| **One-off and continuing training** |  |  |  |
| Ever received formal eCHIS training | Yes | 324 | 94.5 (91.5-96.4) |
|  | No | 19 | 5.5 (3.6-8.5) |
| Refresher training frequency | Monthly | 144 | 42.0 (36.9-47.3) |
|  | Quarterly | 49 | 14.3 (11.0-18.4) |
|  | Annually | 85 | 24.8 (20.5-29.6) |
|  | Never | 65 | 19.0 (15.2-23.4) |
| Mentorship programme available | Yes | 211 | 61.5 (56.3-66.5) |
|  | No | 117 | 34.1 (29.3-39.3) |
|  | Not sure | 15 | 4.4 (2.7-7.1) |
| Trained on data privacy | Yes | 252 | 73.5 (68.6-77.9) |
|  | No | 81 | 23.6 (19.4-28.4) |
|  | Not sure | 10 | 2.9 (1.6-5.3) |
| **User capability** |  |  |  |
| Digital comfort | Very comfortable | 199 | 58.0 (52.7-63.1) |
|  | Somewhat comfortable | 128 | 37.3 (32.4-42.6) |
|  | Not comfortable | 16 | 4.7 (2.9-7.4) |
| Prior digital experience | Yes | 179 | 52.2 (46.9-57.4) |
|  | No | 141 | 41.1 (36.0-46.4) |
|  | Not sure | 23 | 6.7 (4.5-9.9) |
| **Technical support** |  |  |  |
| Availability of technical support | Excellent | 51 | 14.9 (11.5-19.0) |
|  | Good | 98 | 28.6 (24.0-33.6) |
|  | Fair | 114 | 33.2 (28.5-38.4) |
|  | Poor | 38 | 11.1 (8.2-14.8) |
|  | Not available | 42 | 12.2 (9.2-16.1) |
| Response time when support was needed | Immediately | 69 | 20.1 (16.2-24.7) |
|  | Within a day | 78 | 22.7 (18.6-27.5) |
|  | More than two days | 110 | 32.1 (27.4-37.2) |
|  | No support received | 86 | 25.1 (20.8-29.9) |
| **System performance and use** |  |  |  |
| System crash frequency | Never | 25 | 7.3 (5.0-10.5) |
|  | Once | 78 | 22.7 (18.6-27.5) |
|  | 2-3 times | 83 | 24.2 (20.0-29.0) |
|  | More than 3 times | 157 | 45.8 (40.6-51.1) |
| Ease of access | Very easy | 142 | 41.4 (36.3-46.7) |
|  | Somewhat easy | 149 | 43.4 (38.3-48.7) |
|  | Difficult | 34 | 9.9 (7.2-13.5) |
|  | Very difficult | 18 | 5.2 (3.3-8.1) |
| Frequency of eCHIS use | Daily | 148 | 43.1 (38.0-48.4) |
|  | Weekly | 19 | 5.5 (3.6-8.5) |
|  | Monthly | 137 | 39.9 (34.9-45.2) |
|  | Rarely | 39 | 11.4 (8.4-15.2) |
| **Outcomes** |  |  |  |
| System works consistently | Yes | 97 | 28.3 (23.8-33.3) |
|  | No | 216 | 63.0 (57.7-67.9) |
|  | Not sure | 30 | 8.7 (6.2-12.2) |
| Satisfaction with eCHIS | Very satisfied | 71 | 20.7 (16.7-25.3) |
|  | Satisfied | 118 | 34.4 (29.6-39.6) |
|  | Neutral | 116 | 33.8 (29.0-39.0) |
|  | Dissatisfied | 22 | 6.4 (4.3-9.5) |
|  | Very dissatisfied | 16 | 4.7 (2.9-7.4) |
| **Workforce** |  |  |  |
| Staff have left because of eCHIS | Yes | 67 | 19.5 (15.7-24.1) |
|  | No | 248 | 72.3 (67.3-76.8) |
|  | Not sure | 28 | 8.2 (5.7-11.5) |
| Staff retention challenge | Yes | 116 | 33.8 (29.0-39.0) |
|  | No | 208 | 60.6 (55.4-65.7) |
|  | Not sure | 19 | 5.5 (3.6-8.5) |

Percentages use all 343 respondents as the denominator. CI, Wilson score confidence interval.

### 3.3 Determinants of System Performance for Users

In bivariate analysis, the ever-trained indicator showed no association with satisfaction (ρ = −0.03, p = 0.56) or perceived consistency (ρ = 0.01, p = 0.89). Every continuing-support and system-performance variable was associated with both outcomes. The strongest associations were technical support response time with consistency (ρ = 0.37, p < 0.001), and ease of access with crash frequency (ρ = −0.36) and satisfaction (both p < 0.001).

The hierarchical models confirmed this pattern (Table 3). Adding one-off training to the capability block did not improve either model (satisfaction: likelihood-ratio χ² = 0.22, df = 1, p = 0.64; consistency: χ² = 0.10, df = 1, p = 0.75). Adding the continuing-support block improved both (satisfaction: χ² = 21.3, df = 3, p < 0.001; consistency: χ² = 35.5, df = 3, p < 0.001). For consistency, McFadden’s pseudo-R² rose from 0.06 to 0.16. Adding system performance further improved the satisfaction model (χ² = 30.2, df = 2, p < 0.001) but not the consistency model (χ² = 1.9, df = 2, p = 0.39).

**Table 3.** Determinants of Satisfaction and Perceived System Consistency: Full Hierarchical Models.

| Variable | Satisfaction<br>aOR (95% CI) | p | Consistency<br>aOR (95% CI) | p |
|---|---|---|---|---|
| **User capability** |  |  |  |  |
| Digital comfort (per level) | 1.98 (1.07-3.66) | 0.030 | 2.01 (1.25-3.24) | 0.004 |
| Prior digital experience | 1.24 (0.72-2.11) | 0.438 | 0.75 (0.40-1.39) | 0.361 |
| >6 years in community health | 1.20 (0.75-1.92) | 0.447 | 1.10 (0.44-2.71) | 0.840 |
| **One-off training** |  |  |  |  |
| Formal eCHIS training (ever) | 0.78 (0.28-2.19) | 0.642 | 0.76 (0.24-2.41) | 0.646 |
| **Continuing support** |  |  |  |  |
| Refresher training frequency (per level) | 1.20 (0.95-1.53) | 0.131 | 1.25 (0.94-1.67) | 0.131 |
| Mentorship available | 0.75 (0.39-1.45) | 0.397 | 1.52 (0.64-3.59) | 0.339 |
| Technical support speed (per level) | 1.34 (0.93-1.93) | 0.118 | 1.87 (1.35-2.58) | <0.001 |
| **System performance** |  |  |  |  |
| Crash frequency (per level) | 0.60 (0.43-0.84) | 0.003 | 0.83 (0.61-1.14) | 0.260 |
| Ease of access (per level) | 1.58 (1.04-2.42) | 0.033 | 1.12 (0.77-1.63) | 0.562 |
| **Contribution of each block (likelihood-ratio test; McFadden pseudo-R² after entry)** |  |  |  |  |
| Capability only | Reference | R² = 0.054 | Reference | R² = 0.062 |
| + One-off training | χ² = 0.2, df = 1 | 0.64; R² = 0.054 | χ² = 0.1, df = 1 | 0.75; R² = 0.062 |
| + Continuing support | χ² = 21.3, df = 3 | <0.001; R² = 0.079 | χ² = 35.5, df = 3 | <0.001; R² = 0.164 |
| + System performance | χ² = 30.2, df = 2 | <0.001; R² = 0.115 | χ² = 1.9, df = 2 | 0.39; R² = 0.169 |

Satisfaction: proportional-odds ordinal logistic regression (n = 307). Consistency: binary logistic regression, 'not sure' excluded (n = 281). Standard errors clustered on 22 health facilities. Ordered predictors entered as linear scores (odds ratio per one-level increase). aOR, adjusted odds ratio; CI, confidence interval; df, degrees of freedom.

In the full models, crash frequency was the strongest independent correlate of satisfaction (adjusted odds ratio [aOR] 0.60 per level, 95% CI 0.43-0.84), followed by ease of access (aOR 1.58, 1.04-2.42) **(Fig 4).** Technical support response time was the strongest correlate of perceived consistency (aOR 1.87, 1.35-2.58). Average predicted probabilities of reporting a consistent system rose from 15% among respondents who received no support to 50% among those helped immediately (**Fig 3A).** The share of respondents who were satisfied fell from 95% among those who had never experienced a crash to 41% among those who had experienced more than three (**Fig 3B).** Capability still mattered. Digital comfort was independently associated with both outcomes (aOR 1.98, 1.07-3.66 and 2.01, 1.25-3.24). However, its odds ratio fell by a third or more once support and system variables were added, from 3.04 to 1.98 for satisfaction and from 3.42 to 2.01 for consistency. Neither refresher training frequency nor mentorship was independently significant once technical support and system performance were accounted for. Both were positively associated with the outcomes in bivariate analysis and correlated with each other (ρ = 0.43).

**Fig 3.**
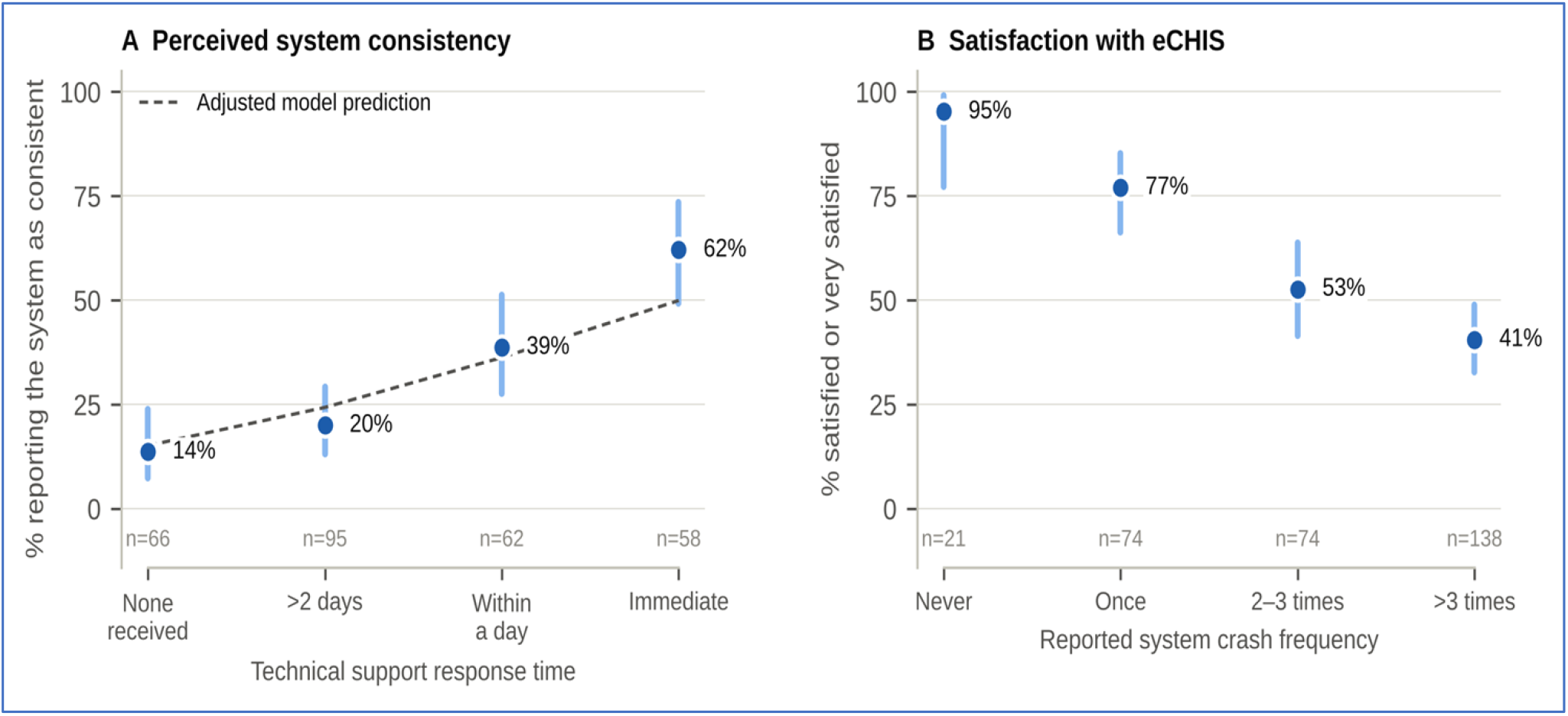
Gradients in user-reported system performance. (A) Percentage of respondents reporting the system as consistent, by technical support response time, with average predicted probabilities from the adjusted logistic model (dashed line); ‘not sure’ responses excluded (n = 281). (B) Percentage satisfied or very satisfied, by reported crash frequency (n = 307). Vertical lines show Wilson 95% confidence intervals.

**Fig 4.**
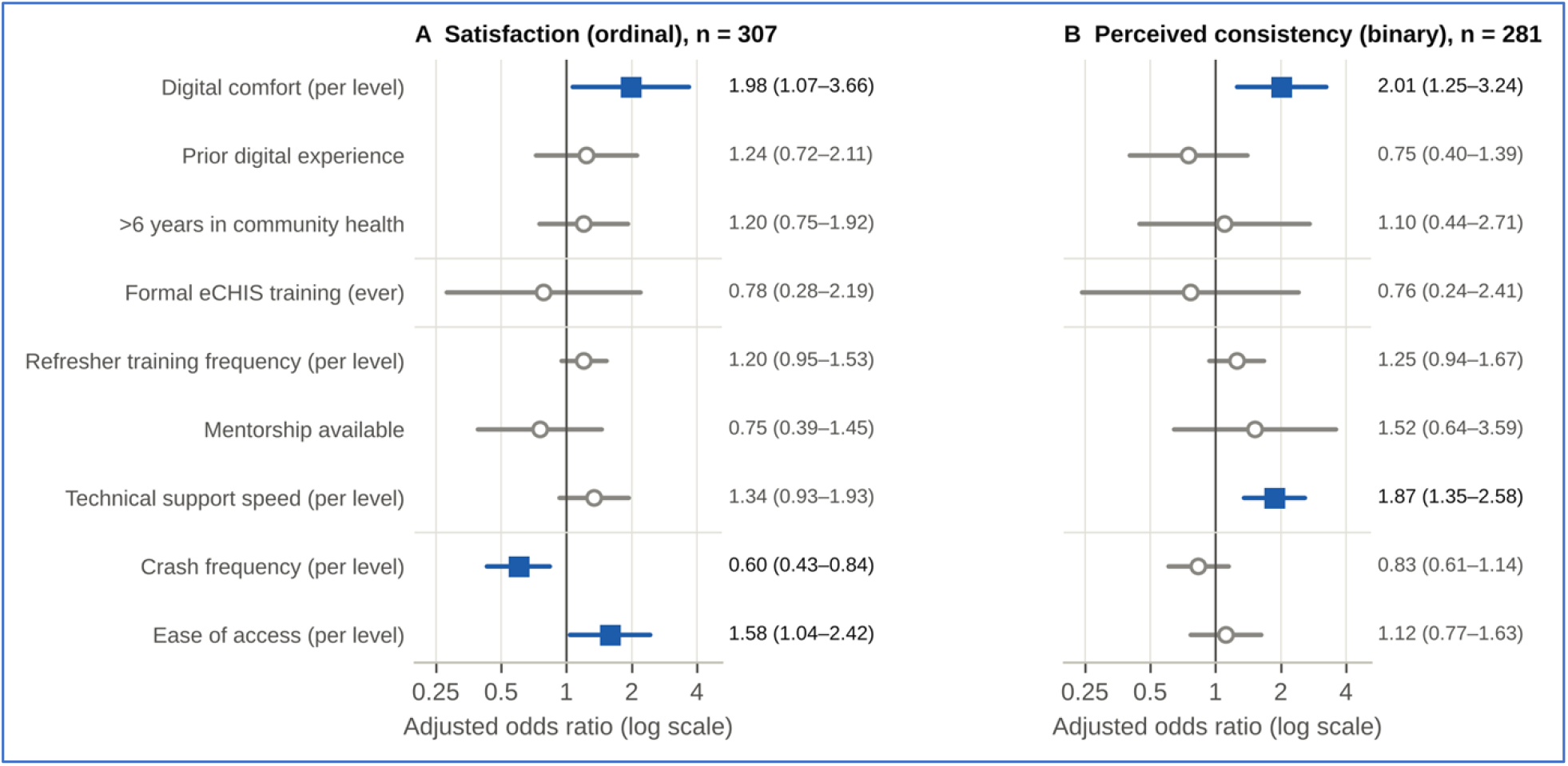
Adjusted odds ratios from the full hierarchical models. (A) Satisfaction, proportional-odds ordinal logistic regression. (B) Perceived consistency, binary logistic regression. Filled squares mark p < 0.05. Horizontal lines show 95% confidence intervals with standard errors clustered on health facility.

These findings held across sensitivity analyses (S2 Table). Restricting the sample to CHPs left the estimates for crashes, access, technical support and digital comfort essentially unchanged. Recoding “not sure” responses as “no” also made little difference. Using technical support availability instead of response time produced a significant association with both satisfaction (aOR 1.52, p < 0.001) and consistency (aOR 1.37, p = 0.03). With satisfaction dichotomised, crash frequency (aOR 0.57, 0.40-0.81) and ease of access (aOR 1.73, 1.05-2.87) remained significant. Cut-point-specific models gave consistent estimates for crash frequency across thresholds. Estimates at the lowest threshold were imprecise because only 38 respondents fell below neutral. Formal training had no significant association with either outcome in any specification.

### 3.4 Where reporting quality is secured and where it is lost

The observation checklist showed the same split between capture and what follows (Fig 5, Table 4). At capture, the application’s design largely ensured quality. All required fields were completed at every site, and observers noted that a form “if not filled [allows] no submission”. No entries were skipped at four of five sites, and entry formats were standardised at four of five.

**Fig 5.**
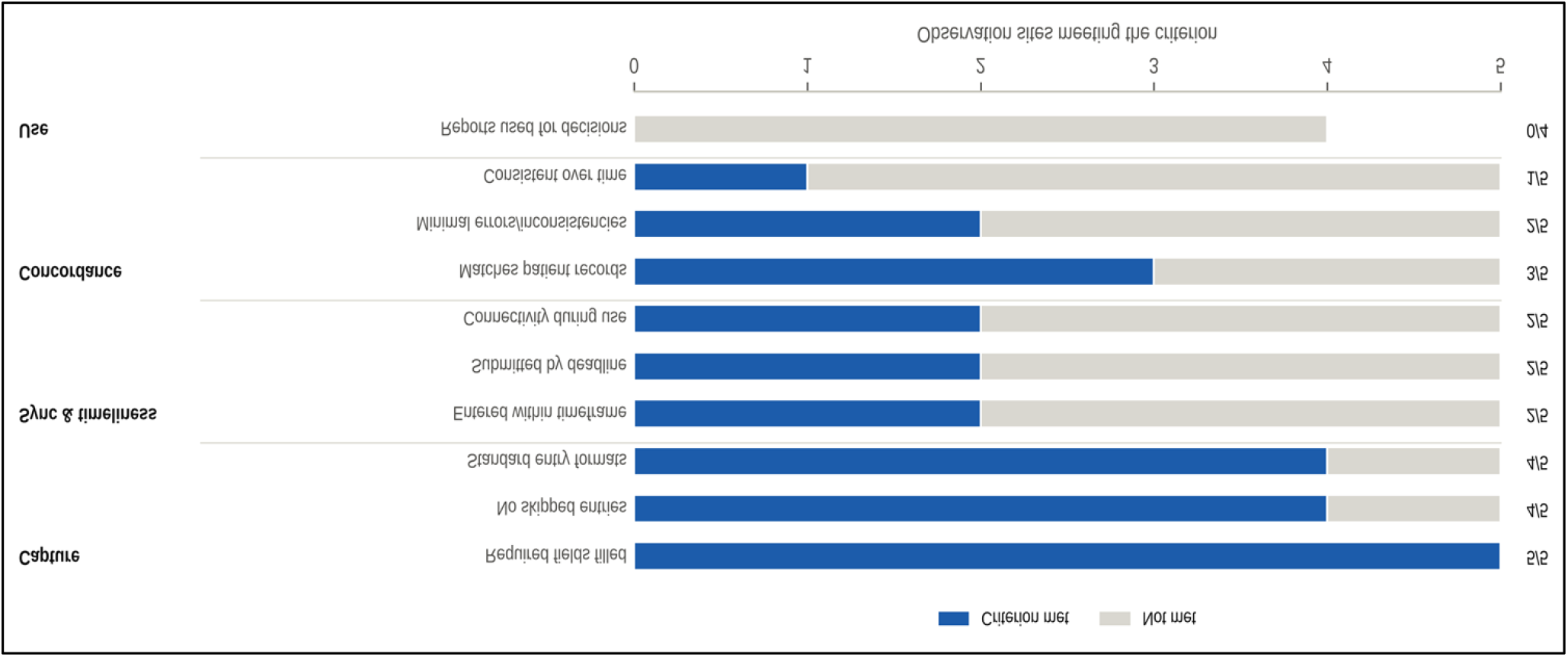
Reporting quality along the reporting chain at five observation sites. Number of sites meeting each checklist criterion, grouped by stage of the reporting chain. The use indicator was recorded at four sites.

**Table 4.** Observation checklist: selected indicators of reporting quality and support at five sites.

| **Checklist indicator** | **Sites meeting criterion** | **Illustrative observer notes** |
|---|---|---|
| **Capture** |  |  |
| All required data fields are filled | 5/5 | “If not filled no submission” |
| No missing or skipped data entries | 4/5 |  |
| Data entry formats are standardized across users | 4/5 | “Duplication of one client many times” |
| **Transmission** |  |  |
| Data entered immediately or within recommended timeframe | 2/5 | “Synchronization is … main challenge”; “system failure” |
| Reports submitted within required deadlines | 2/5 | “By use of own resources”; “difficulty in synchronization” |
| **Concordance** |  |  |
| Data entered into eCHIS matches patient records | 3/5 | “Missing patients, duplication of patients” |
| Reports from eCHIS match manual records (if available) | 3/5 | “Due to delayed syncing data not up to date” |
| Data retrieved from eCHIS is consistent over time | 1/5 | “Difference between hard and soft copy” |
| **Use** |  |  |
| Reports used for decision-making | 0/4 | “Does not portray the clear picture on the ground” |
| **Support** |  |  |
| Prompt response to system-related issues | 2/5 | “If it crashes they wait until the system is back”; “no support at all” |
| Availability of user manuals, help guides, or online support | 0/5 |  |
| Internet connectivity available during system use | 2/5 | “Offline, use of their own resources when syncing” |
| Presence of uninterrupted power supply (UPS) or generator | 0/5 |  |
| Regular system maintenance is conducted | 1/5 |  |
| Evidence of recent eCHIS training (within last 6 months) | 2/5 | “Training was done 2 years ago during implementation” |

Further along the chain, quality fell away. Data were entered within the recommended timeframe at two of five sites and reports submitted by the deadline at two of five. Observers attributed the failures to “difficulty in synchronization” and “system failure”. At one site where reports arrived on time, this was “by use of own resources”. Records matched patient registers at three of five sites, and data were consistent over time at only one. Observers recorded duplicated clients, patients who disappeared from the system after failed synchronisation, and discontinued clients who still appeared as active. At none of the four sites assessed were eCHIS reports used for decisions. One observer noted that the reports “does not portray the clear picture on the ground”.

The support environment observed at the sites matched these breakdowns. None of the five sites had user manuals or help guides. None had an uninterrupted power supply or generator. One had an alternative power source, one had backup devices and one had regular system maintenance. Two sites had connectivity during use. Recent training (within six months) was evident at two sites. At one site, observers noted that training “was done 2 years ago during implementation”.

### 3.5 Who keeps the system running

Responses to the multiple-response items show where users located the problem (Table 5). Poor connectivity was the most frequently reported usage challenge (75.5%), followed by system downtime or slowness (51.0%). Inadequate training was cited by 18.1%. In their free-text recommendations, respondents most often asked for data bundles (46.2% of those who answered), stable internet (39.9%), more or refresher training (32.7%), better-quality replacement phones (22.0%) and payment of the county stipend (15.4%). Across all three items, 43.4% of respondents cited the cost of data bundles, and 47.8% cited either bundles or stipends. Users also wanted training. Availability of training was the improvement area selected most often (63.3%), and online tutorials were the preferred format (49.9%). This preference was for continuing, accessible learning rather than another round of initial instruction.

**Table 5.** Problems and remedies raised in multiple-response survey items, n (%).

| **Category** | **Usage challenges (n = 343)** | **Areas to improve (n = 343)** | **Recommendations (n = 318)** |
|---|---|---|---|
| Connectivity / network | 259 (75.5) | - | 127 (39.9) |
| System downtime / speed / hanging | 175 (51.0) | 83 (24.2) | 8 (2.5) |
| Data bundles (airtime) costs | 14 (4.1) | 1 (0.3) | 147 (46.2) |
| Stipend / remuneration | 11 (3.2) | - | 49 (15.4) |
| Device quality / replacement | 1 (0.3) | - | 70 (22.0) |
| Training | 62 (18.1) | 217 (63.3) | 104 (32.7) |
| Integration with other systems | 1 (0.3) | 176 (51.3) | 6 (1.9) |
| Interface / repeated questions | - | 105 (30.6) | 4 (1.3) |
| Workload / households per CHP | - | - | 9 (2.8) |
| Technical support | - | - | 5 (1.6) |
| **Preferred format for future training** |  |  |  |
| Online tutorials |  |  | 171 (49.9) |
| Formal workshops |  |  | 96 (28.0) |
| On-the-job practice |  |  | 44 (12.8) |
| Peer mentorship |  |  | 31 (9.0) |

Responses were coded with a keyword dictionary; a response could fall in more than one category. Across the three items, 149 respondents (43.4%) raised data bundle costs and 164 (47.8%) raised bundles or stipends. Preferred training format is reported for all 343 respondents (one missing).

Key informants described a support system that relied on frontline workers to fill its gaps. Ten of 13 reported either no technical support or only troubleshooting by CHAs and focal persons, and nine described support as inadequate or absent. When the application failed, the most common response was for users to uninstall and reinstall it themselves (6 of 13). Four informants described waiting with no action or long delays. Nine described internet access as unreliable, and one explained that connectivity “depends on data bundles”. Bundles were mentioned by 10 of the 13 informants. Six referred to “whitelisting” CHP phone lines so that eCHIS traffic carries no data charge, either as a government measure or as one still needed. Seven informants judged the data inconsistent and seven judged them unreliable. However, nine believed eCHIS had improved accuracy at the point of capture through “real time reports at service point” and the absence of “guesswork”. Ten reported that training had happened once or infrequently. Observers recorded the human side of this arrangement: “most of the CHPs lack data bundles”, “reduced morale due to use of their own resources” and “reduced morale due to delayed monthly stipend”.

### 3.6 Integration

Table 6 brings the three strands together. They converged on four points. One-off training was near universal and did not explain differences in how the system performed. Continuing support, especially technical response, did. Completeness was secured at capture, while timeliness, consistency and use failed at the synchronisation stage. The recurrent costs of connectivity fell on CHPs. The main divergence concerned accuracy. Informants credited eCHIS with more accurate capture, while observers found records matching source registers at only three of five sites. This difference is consistent with the chain model: data entered accurately can still arrive late, duplicated or incomplete once synchronisation fails.

**Table 6.** Joint display integrating survey, observation and key informant findings.

| Theme | Survey (n = 343) | Observation (5 sites) | Key informants (n = 13) | Integrated inference |
|---|---|---|---|---|
| One-off training does not explain performance | 94.5% formally trained; no association with satisfaction or consistency in any model | Recent training at 2/5 sites; “training was done 2 years ago” | Training once or infrequent (10/13) | Convergent: initial training is near universal and not the differentiating factor |
| Continuing support matters | 57.1% had delayed or no support; support response time aOR 1.87 for consistency | Prompt response to failures at 2/5 sites; no manuals at any site | No or peer-only support (10/13); self-reinstallation of the app (6/13) | Convergent: frontline workers fill the support gap themselves |
| Quality is secured at capture | Ease of access high (84.8% easy) | Required fields filled at 5/5 sites | Improved accuracy at capture (9/13) | Convergent: form logic secures completeness |
| Quality is lost in transmission and use | 70.0% had ≥2 crashes; 63.0% found the system inconsistent | Timely entry 2/5; consistent over time 1/5; used for decisions 0/4 | Data inconsistent (7/13) or unreliable (7/13); missing or duplicated clients (5/13) | Convergent: synchronisation is where quality fails |
| Costs fall on CHPs | 43.4% raised data bundle costs; 46.2% of recommendations asked for bundles | “Use of their own resources”; “CHPs lack data bundles”; reduced morale | Bundles cited by 10/13; “whitelisting” CHP lines cited by 6/13 | Convergent: timeliness depends on private spending by volunteers |
| Accuracy | Not measured directly | Records matched patient registers at 3/5 sites | Accuracy improved (9/13) vs not improved (4/13) | Divergent: accurate capture does not guarantee accurate central records |

aOR, adjusted odds ratio; CHP, community health promoter.

## 4. Discussion

Nearly all eCHIS users in Kisii County had been trained, were comfortable with digital tools and could use the application. Nevertheless, they worked with a system most experienced as inconsistent and produced reports that no one at the observed sites used for decisions. How well the system worked for users was not associated with whether they had been trained. It was associated with how quickly help arrived when the system failed, how often it crashed and how easily it could be reached. Observations and interviews located the loss of quality after the point of capture: at synchronisation, which depended on connectivity that CHPs often paid for themselves. Data quality in this system came less from what users knew than from the support ecology around them.

These findings fit, and sharpen, the international evidence on training. Reviews and trials show that training builds knowledge and skill but rarely improves routine data quality on its own, and that successful interventions combine training with supervision, mentorship and feedback ^13,15,16,45^. In our data, initial formal training reached almost everyone and made no measurable difference. By contrast, the continuing-support block explained a substantial share of the variation in both outcomes. Within that block, technical response time carried most of the independent association. Refresher training and mentorship were positively associated with the outcomes in bivariate analysis, but they were correlated with each other and with technical support. This pattern suggests that facilities providing one form of continuing support tend to provide the others. It agrees with Ethiopian accounts that training without follow-up leaves confidence gaps ^9,10^. It also agrees with Kenyan reports that short trainings are inadequate ^6 6^. Users in our study wanted more training, but they wanted it continuing and accessible, mostly as online tutorials, which a single induction course cannot provide.

Digital comfort remained independently associated with both outcomes, so individual capability is not irrelevant. However, its effect fell by a third or more once support and system variables were added. This is consistent with the idea that users who feel competent are partly those who are well supported. Competency and support are therefore better understood as joint products of the same environment than as rival explanations. This matters for policy because a competency framework that certifies individual skill will not, by itself, change the conditions in which those skills are used.

The observation data clarify why training and competence are insufficient. The eCHIS application enforced completeness by design: a form could not be submitted until required fields were filled. Completeness, the dimension most often reported as improved by digitisation ^1,3^, was therefore secured at every site. The dimensions that depend on infrastructure were not. Timeliness, concordance with source registers, consistency over time and use all failed at most sites, and observers and informants linked these failures to synchronisation. The same sequence has been described in Ethiopia, Uganda and South Africa. There, connectivity and synchronisation failures delayed uploads and produced duplicated or missing records, and integrated data lagged days behind capture ^20–22,24^. Our findings add that a single headline indicator such as completeness can conceal these failures. A county’s reported completeness may look good while the data that reach supervisors are late, duplicated and not trusted for decisions. In PRISM terms, the technical determinants of the system set a ceiling on performance that behavioural interventions cannot raise ^35,36 35,36^. In design-reality terms, the application assumed a connected, powered and financed environment that frequently did not exist ^37,39^.

The third finding concerns who pays. Almost half of respondents raised the cost of data bundles or unpaid stipends without being asked about costs. Informants described CHPs using their own resources to synchronise and repairing the application themselves by reinstalling it. Kisii is not unusual in this. Across low- and middle-income countries, programmes typically finance devices and initial training while the recurrent costs of airtime, data, charging and repair shift to workers^28,31,46 28,31,46.29^ In Tanzania, frontline workers have reported spending more on work data than they earned. Our data cannot show whether these costs affected retention, although a third of respondents (33.8%) reported staff retention challenges. They do show that the timeliness of reporting partly depended on whether volunteers could afford to upload. Elsewhere, cost-shifting and payment delays have been linked to lower motivation and performance, and digital programmes that covered these costs achieved better retention^32–34,47^. When a public reporting system depends on private spending by its least-paid workers, its data quality inherits the instability of their finances.

These findings have practical implications for Kenya’s eCHIS and for similar systems elsewhere. The first concerns financing. eCHIS connectivity should be treated as a core programme cost, not a personal expense. Zero-rating or “whitelisting” eCHIS traffic, which informants proposed, is one way to do this, and direct data reimbursement is another. The second concerns technical support. Support should operate to defined service standards, including maximum response times, a reachable help desk, routine maintenance and replacement devices. Response time was the factor most strongly associated with perceived consistency in our data. The third concerns monitoring. Synchronisation should be monitored as a data quality indicator in its own right. Lag between capture and upload, failed synchronisations and duplicate records can be measured from system logs, and these measures would show the failures that completeness indicators conceal^21,26^. The fourth concerns training. It should be reorganised as continuing support, delivered through accessible online materials, refresher sessions and mentorship linked to supervision, rather than as a one-time event ^15,16^. A national eCHIS competency framework would be strengthened by including these support standards alongside individual competencies.

### 4.1 Strengths and limitations

This study combined a large survey of eCHIS users with direct observation and key informant interviews in a single county. All estimates were calculated from record-level data with clustered standard errors and several sensitivity analyses. The study also has limitations of scope. First, the design was cross-sectional, so the associations cannot establish direction. For example, dissatisfied users may judge support more harshly. Second, the survey outcomes reflect how users experienced the system. They are not audited measures of data quality, and evidence on data quality itself comes from five observation sites and 13 interviews. Third, the observation sites were identified at sub-county level and selected purposively, so their results describe conditions at those sites and should not be generalised as county-wide prevalence. Fourth, key informant responses were recorded as structured summaries rather than verbatim transcripts, which limits the depth of the qualitative analysis. Fifth, each construct was measured with single items, and some multiple-response items offered preset options, which may have shaped the categories respondents chose. Sixth, the sample was dominated by experienced CHPs, so new users are underrepresented. Finally, the study covers one county in the second year of implementation. Future work should link worker surveys to system logs of synchronisation lag and duplication, compare counties with different financing arrangements, and prospectively test support standards.

### 4.2 Conclusion

Training put eCHIS into the hands of almost every community health promoter in Kisii County. However, it was not what determined whether the system delivered timely, consistent and usable data. Those outcomes depended on continuing support, system stability and connectivity, much of which CHPs financed themselves. Efforts to improve community health data quality in Kenya should finance connectivity, responsive technical support, and synchronisation monitoring with the same priority as competency development.

## Data Availability

We, the authors, are committed to providing access to and making available research data for reviewers' scrutiny and validation to fulfill the ethical requirement of transparency and openness in the research process.

## Acknowledgments

We thank the community health promoters, community health assistants and focal persons of Kisii County who took part in the study, and the Kisii County Department of Health for facilitating access.

## Supporting information

**S1 Table. Observation checklist results by indicator.** Number of sites meeting each of the 40 checklist criteria, with observers’ comments.

**S2 Table. Sensitivity analyses.** Adjusted odds ratios from the full models under four alternative specifications.

**S3 Table. Block-wise model estimates.** Adjusted odds ratios for each predictor at each step of the hierarchical models.

**S1 Checklist. STROBE checklist.** Completed STROBE checklist for cross-sectional studies.

